# Multimodal neuroimaging and suicidality in a US population-based sample of school-aged children

**DOI:** 10.1101/19013193

**Authors:** Pablo Vidal-Ribas, Delfina Janiri, Gaelle E. Doucet, Narun Pornpattananangkul, Dylan M. Nielson, Sophia Frangou, Argyris Stringaris

## Abstract

**Importance:** Suicide deaths and suicidality are considered a public health emergency, yet their brain underpinnings remain elusive.

**Objective:** To examine individual, environmental, and clinical characteristics, as well as multimodal brain imaging correlates of suicidality in a US population-based sample of school-aged children.

**Design:** Cross-sectional analysis of the first wave of data from the Adolescent Brain Cognitive Development study

**Setting:** Multicenter population-based study

**Participants:** Children aged 9-10 years from unreferred, community samples with suicidality data available (n=7,994). Following quality control, we examined structural magnetic resonance imaging (sMRI) (n=6,238), resting state functional MRI (rs-fMRI) (n=4,134), and task-based fMRI (range n=4,075 to 4,608).

**Exposure:** Lifetime suicidality, defined as suicidal ideation, plans and attempts reported by children or/and caregivers.

**Main Outcomes and Measures:** Multimodal neuroimaging analyses examined differences with Welch’s t-test and Equivalence Tests, with observed effect sizes (ES, Cohen’s *d*) and their 90% confidence interval (CI) < |0.15|. Predictive values were examined using the area under precision-recall curves (AUPRC). Measures included, cortical volume and thickness, large-scale network connectivity and task-based MRI of reward processing, inhibitory control and working memory.

**Results:** Among the 7,994 unrelated children (3,757 females [47.0%]), those will lifetime suicidality based on children (n=684 [8.6%]; 276 females [40.4%]), caregiver (n=654 [8.2%]; 233 females [35.6%]) or concordant reports (n=198 [2.5%]; 67 females [33.8%]), presented higher levels of social adversity and psychopathology on themselves and their caregivers compared to never-suicidal children (n=6,854 [85.7%]; 3,315 females [48.3%]). A wide range of brain areas was associated with suicidality, but only one test (0.06%) survived statistical correction: children with caregiver-reported suicidality had a thinner left bank of the superior temporal sulcus compared to never-suicidal children (ES=-0.17, 95%CI -0.26, -0.08, *p*_FDR_=0.019). Based on the prespecified bounds of |0.15|, ∼48% of the group mean differences for child-reported suicidality comparisons and a ∼22% for parent-reported suicidality comparisons were considered equivalent. All observed ES were relatively small (*d*≤|0.20|) and with low predictive value (AUPRC≤0.10).

**Conclusion and Relevance:** Using commonly-applied neuroimaging measures, we were unable to find a discrete brain signature related to suicidality in youth. There is a great need for improved approaches to the neurobiology of suicide.

## Introduction

Rates of suicide deaths and suicidality —defined as suicide ideation, plans and attempts - have risen over 50% amongst young people in the last decade ^1-3^, making suicide the second-leading cause of death in those aged 10-19 years ^2,4^. Whereas individual, environmental and clinical risk factors for suicidality have been well-established ^5-14^, these have demonstrated low predictive validity ^15-17^. In response, the number of studies examining neurobiological underpinnings of suicidality has grown exponentially in the last two decades ^18^. Nevertheless, our understanding and utility of the neural mechanisms underlying suicidality is still poor, especially in young children, for several reasons.

First, it is still unclear whether findings from neuroimaging studies examining suicidality apply to children, since most studies have been conducted in adult samples. Second, results of these studies have been inconsistent. Whereas systematic reviews on the topic suggest that suicidality is associated with abnormalities in regions involved in affective processing and impulsive regulation, the specific regions highlighted in each review differ, and all emphasize the modest sample sizes, heterogeneity, and lack of replicability across studies ^18-21^. In addition, meta-analyses of structural and functional imaging studies have failed to find differences between suicidal and non-suicidal participants ^22-24^, and those that found differences were either based on a small number of studies or reported inconsistent findings ^24,25^. Third, it is unclear whether the effect sizes (ES) of any described neural correlates of suicidality are large enough to have clinical utility. Studies with small sample sizes have limited power to detect differences ^19^. However, finding no difference does not mean that the difference equals zero; the observed ES could be considered large enough to be meaningful. On the other hand, studies with large sample sizes are more powered to detect small differences; yet, the observed ES of such differences might be too small for practical purposes ^23^. To examine whether an observed ES is large enough to be considered meaningful one can test for equivalence ^26^, an approach originally employed in the field of pharmacokinetics ^27^ with the aim of showing that a new cheaper drug was practically as effective as an existing one.

In the current study, we employed data from a large population-based sample from the Adolescent Brain and Cognitive Development (ABCD) study (https://abcdstudy.org/) ^28,29^ to examine the correlates of suicidal behaviors using a multi-informant approach. In children aged 9-10 years, we first examined individual, environmental and clinical correlates of suicidality typically found at this age ^9-11,13,14,30^. Next, we sought to identify associations between suicidality and brain morphometry, functional connectivity at rest, and functional measures during three tasks involving reward processing ^31-33^, inhibitory control ^34,35^, working memory ^36,37^ and affective processing ^38,39^. We tested for differences in these measures using a traditional null hypothesis significance test, and complemented our analyses with Equivalence testing ^26^ to examine whether observed ES were large enough to be considered meaningful based on a prespecified benchmark. Finally, we examined the ability of neural correlates to predict suicide cases in our sample.

## Methods

### The ABCD study

All the data used here were accessed from the ABCD Study Curated Annual Release 2.1 and are available on request from the NIMH Data Archive (https://data-archive.nimh.nih.gov/abcd). The baseline ABCD sample consists of 11,875 children from 22 sites across the United States that match the demographic profile of the American Community Survey ^29^. The University of California at San Diego Institutional Review Board was responsible for the ethical oversight of the ABCD study. The present study is based on 7,994 unrelated ABCD participants for whom complete self-report *and* caregiver data on childhood suicidality were available. As detailed in the Supplementary **eMethods** and illustrated in **eFigure 1**, the neuroimaging analyses involved subsamples based on the availability of high-quality magnetic resonance imaging (MRI) data for each modality.

**Figure 1.**
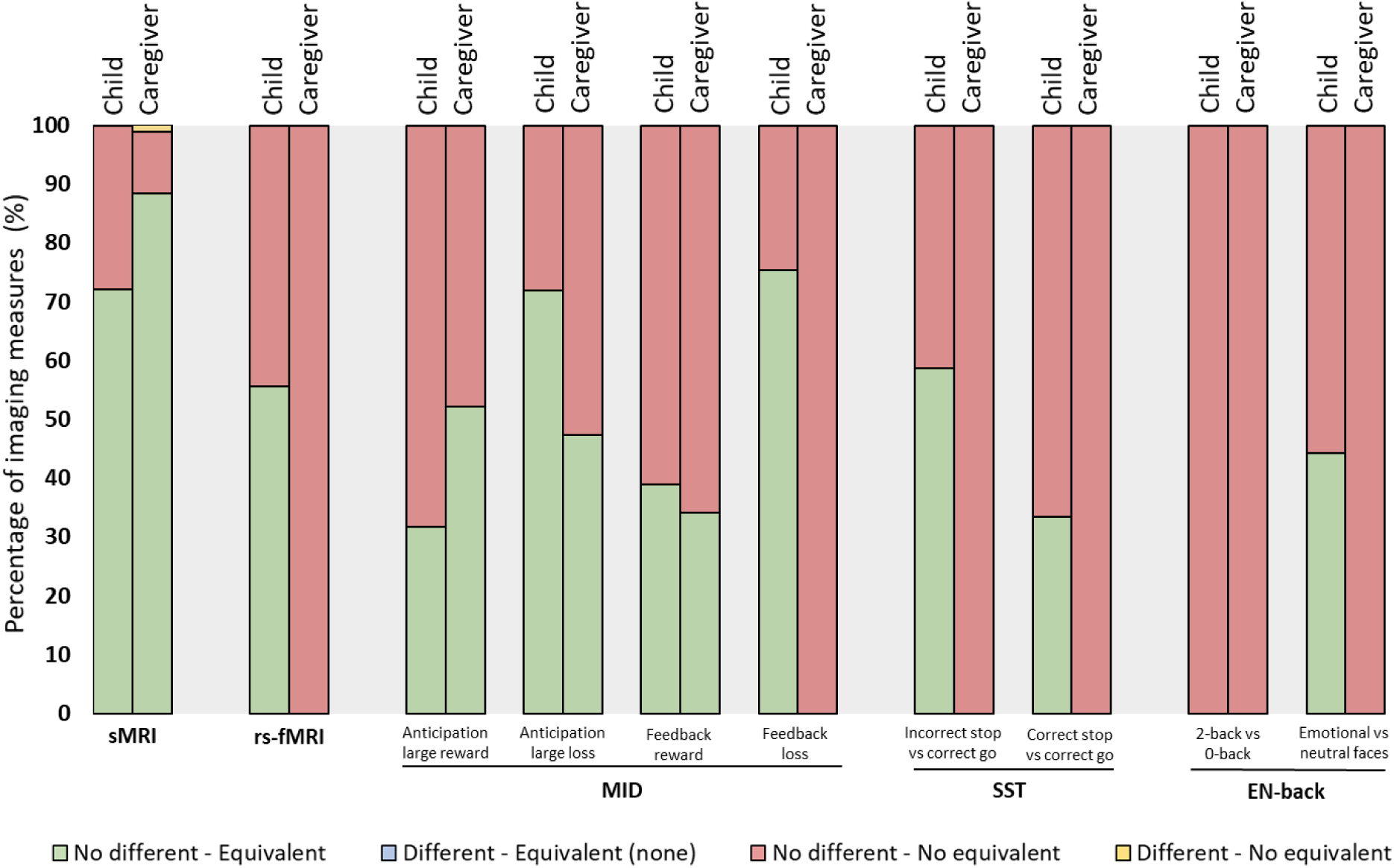
Percentage of outcomes of the Welch’s t-test and Equivalence test for each imaging modality by suicidality group comparison. For each informant, structural MRI examined 86 regions, resting-state fMRI examined 306 connectivity indices, and task-based fMRI examined activations in 167 regions. No evidence of difference (Welch’s t-test, p_FDR_ ≥0.05, 95% confidence interval (CI) includes zero), Evidence of difference (Welch’s t-test, p_FDR_<0.05, 95%CI does not include zero). Evidence of equivalence (Equivalence test, p_FDR_<0.05, 90%CI does not overlap with bounds); No evidence of equivalence (Equivalence test, p_FDR_≥0.05, 90%CI overlaps with bound/s). sMRI, structural MRI. rs-fMRI, resting-state fMRI. MID, Monetary incentive delay task. SST, Stop signal task. EN-back, Emotional n-back task.

### Determination of childhood suicidality

Suicidality in children was assessed using the child- and caregiver-report of the computerized Kiddie Schedule for Affective Disorders and Schizophrenia for DSM-5 (KSADS-5) ^40^. A detailed description of the assessment is provided in the **eMethods**. Based on children and caregiver reports, four suicidality groups were computed: 1) child-reported suicidality 2) caregiver-reported suicidality, 3) concordantly-reported suicidality (i.e., both child and parent endorse at least one item), and 4) never-suicidal (i.e., both child and parent do not endorse any item).

### Individual and environmental characteristics

We examined individual and environmental characteristics, as well as clinical factors of the child and caregivers that have been associated with suicidality in previous studies ^5-7,9-11,13^. A detailed description of the factors and instruments employed to assess these variables are provided in **Supplementary** e**Table 1**.

### Neuroimaging

High-resolution T_1_-weighted images as well as resting-state and task-based fMRI data were obtained at each ABCD site using 3T MRI systems. In the current study we examined cortical thickness (n=68 parcellations) and subcortical volumes (n=18 parcellations), functional connectivity at rest (n=306 connectivity indices) and neural activations (n=167 parcellations) evoked by three tasks: a modified monetary incentive delay task (MID) ^41^, stop signal task (SST) ^42^ and emotional n-back task (EN-back) ^43,44^. To preserve statistical power we analyzed each modality separately rather than selecting only those children that had high-quality data across all three of the imaging modalities (**eFigure 1**). A detailed description of the acquisition protocols, quality control procedures, imaging processing, and analyses of the ABCD study have been published elsewhere ^28,45^ and are summarized in the **eMethods**.

### Statistical analysis

All analyses compared never-suicidal children with those with endorsed suicidality. A detailed rationale and description of the tests can be found in the **eMethods**.

#### Analysis of individual and environmental characteristics

Group differences in psychosocial factors were examined with Welch’s t-tests ^46^, to allow for unequal number of observations, and chi-squared tests. Results were considered significant at *p*<0.05 with False Discovery Rate (FDR) correction for multiple comparisons.

#### Analysis of differences and equivalence of neuroimaging data

We examined *differences* in neuroimaging measures between groups with Welch’s t-tests to account for unequal number of observations. We examined *equivalence* of mean differences (i.e., whether observed ES of mean differences were meaningful effects) with Equivalence tests (which also included Welch’s t-tests to account for unequal number of observations). In equivalence testing, the observed data are statistically compared against *a priori* specified equivalence interval (d), defined by upper (Δ_*U*_) and lower (−Δ_*L*_) equivalence bounds. The aim of equivalence testing is to reject the null hypothesis that the observed ES (Cohen’s *d*) is at least as extreme as a pre-specified smallest effect size of interest (SESOI). We used the “two one-sided tests” (TOST) procedure ^26,47^ implemented with the *TOSTtwo* function from the library *TOSTER* in R. Given the current sample size and previous results in a large sample ^23^, the upper (Δ_*U*_) and lower (−Δ_*L*_) equivalence bounds were specified as a conservative *d*=0.15 and *d*=-0.15 (i.e., SESOI=|0.15|), which correspond to traditional notions of a “small” ES ^48^. That is, ES with 90%CI within [-0.15, 0.15] were considered statistically equivalence (i.e., not meaningful effects). The threshold for statistical significance for both tests was set at *p* <0.05 after applying FDR-correction for multiple comparisons.

#### Analysis of predictive value of neuroimaging data

Predictive value was estimated with the area under precision-recall curves (AUPRC), which provides more accurate information on the performance of a prediction model than the widely used receiver operating-characteristic (ROC) curves in cases where there is an imbalance in the observations between the two classes ^49^. Precision, or positive predictive value, can be defined as how good a model is at predicting true positive cases. Recall, or sensitivity, can be defined as how good a model is at predicting *all* the true positive cases. A perfect model would have an AUPRC of 1, as in ROC; however, whereas in ROC a random classifier would have an AUC close to 0.50, in PRC that value would be close to the prevalence of positive cases in the population, calculated as y=P/(P+N) (e.g. AUPRC=0.10 if prevalence is 10%).

## Results

### Prevalence of suicidality in the sample

The four suicidality groups were composed as follows: child-reported suicidality (n=684, 8.6%), caregiver-reported suicidality (n=654, 8.2%), concordantly-reported suicidality (n=198, 2.5%), and never-suicidal (n=6,854, 85.7%). Based on child reports, suicidal ideation was endorsed by 8.4% of participants, plans were endorsed by 0.9% of participants and attempts were endorsed by 1.3% of participants. Based on care-giver reports, these rates were 8.1%, 0.6%, 0.5% respectively. Among participants with endorsed suicidality, either by the child or the caregiver (n=1,140, 14.3%), there was an agreement of 17.4% (n=198). **eTable 2** shows the rate of suicidal behaviors reported by either the children or the caregivers and the rate of positive agreement for each item.

### Individual and environmental characteristics

**Table 1** shows the descriptive statistics and comparison of individual and environmental characteristics between the suicidality groups. Several variables differed at p_FDR_<0.05 between the three suicidality groups and the never-suicidal group. Specifically, all suicidality groups presented higher rate of males (59.6%-64.4% vs 51.6%, all *p*<0.001), higher exposure to stressful life events (42.8%-50.9% vs 35.2%, all *p*<0.001), more economic problems in the last 12 months (29%-33.8% vs 21.4%, all *p*<0.001), more family conflict, less positive school environment and higher rates and scores in every individual and parental clinical variable examined, including general psychopathology, psychiatric disorders in the child (35.4%-57.6% vs 21.9%, all *p*<0.001), parental use of mental health services (48.5%-64.7% vs 38.5%, all *p*<0.001), parental hospitalization due to mental health problems (14.5%-24.7% vs 7.8%, all *p*<0.001), maternal alcohol and/or substance use during pregnancy (13%-14.1% vs 8.2%, all *p*<0.001), and parental history of depression (41.4%-61.1% vs 29.6%, all *p*<0.001) and suicide attempt or death (10.6%-18.6% vs 4.7%, all *p*<0.001).

**Table 1.** Individual, environmental and clinical characteristics of the suicidality groups by informant.

|  |  |  |  |  |
| --- | --- | --- | --- | --- |
|  | **Child-reported**<br>**Suicidality**<br>**n=684** | **Caregiver-reported**<br>**Suicidality**<br>**n=654** | **Concordantly-reported**<br>**Suicidality**<br>**n=198** | **Never-Suicidal**<br>**n=6854** |
| **Individual characteristics** |  |  |  |  |
| Sex (% male) | 59.6% * | 64.4% * | 66.2% * | 51.6% |
| Race (%) |  |  |  |  |
| White | 59.4% | 63.1% | 62.8% | 62.9% |
| Black/African American | 17.7% | 12.3% * | 13.3% | 16.5% |
| Asian | 2.8% | 2.1% | 2.6% | 2.1% |
| Multiple races | 15.5% * | 15.9% * | 14.8% | 12.1% |
| Other races | 4.6% | 6.5% | 6.6% | 6.3% |
| Ethnicity (%) |  |  |  |  |
| Hispanic/Latino | 21.0% | 21.1% | 21.1% | 23.1% |
| Total intelligence composite, mean (sd) | 100.7 (18.4) | 99.9 (18.1) | 100.3 (17.6) | 101.0 (18.4) |
| Exposure to stressful events (%) | 42.8% * | 50.9% * | 50.8% * | 35.2% |
| Had a common childhood medical condition (%) | 45.5% | 50.2% * | 49.0% | 42.3% |
| Pubertal development (Caregiver report), mean (sd) |  |  |  |  |
| Females | 2.3 (0.9)* | 2.4 (0.9)* | 2.3 (0.8) | 2.2 (0.9) |
| Males | 1.3 (0.6) | 1.4 (0.6) | 1.4 (0.6) | 1.4 (0.6) |
| Pubertal development (Child report), mean (sd) |  |  |  |  |
| Females | 2.5 (0.8)* | 2.4 (0.8)* | 2.5 (0.8) | 2.3 (0.9) |
| Males | 2.1 (0.7)* | 2.0 (0.7) | 2.1 (0.6) | 1.9 (0.8) |
| **Environmental, family and school characteristics** |  |  |  |  |
| Income level mean (sd) | 7.0 (2.4) | 7.0 (2.4) | 7.2 (2.2) | 7.1 (2.5) |
| Economic problems in the last year (%) | 29.0% * | 32.2% * | 33.8% * | 21.4% |
| Level of highest education achieved by primary caregiver, mean (sd) | 16.5 (2.7) | 16.8 (2.5)* | 17.0 (2.3)* | 16.6 (2.9) |
| Primary caregiver living with partner (%) | 69.3% * | 67.8% * | 67.4% | 73.5% |
| Family conflict (Caregiver report), mean (sd) | 2.8 (2.1)* | 3.4 (2.2)* | 3.3 (2.2)* | 2.4 (1.9) |
| Family conflict (Child report), mean (sd) | 2.8 (2.2)* | 2.6 (2.1)* | 3.1 (2.3)* | 1.9 (1.9) |
| Positive school environment, mean (sd) | 18.8 (3.3)* | 19.0 (3.4)* | 18.3 (3.4)* | 20.0 (2.8) |
| **Individual clinical characteristics** |  |  |  |  |
| General psychopathology (CBCL), mean (sd) T-scores | 52.0 (11.6)* | 57.8 (11.1)* | 59.7 (10.7)* | 45.3 (10.8) |
| Current Psychiatric disorders (Caregiver report) (%) |  |  |  |  |
| Any disorder | 35.4% * | 53.5% * | 57.6% * | 21.9% |
| Any depressive disorder | 1.3% * | 1.8% * | 3.5% * | 0.2% |
| Any anxiety disorder | 17.5% * | 25.8% * | 31.3% * | 10.2% |
| ADHD | 17.1% * | 27.5% * | 29.8% * | 9.5% |
| ODD | 10.3% * | 18.8% * | 20.7% * | 4.3% |
| CD | 5.9% * | 13.0% * | 12.6% * | 2.1% |
| PTSD | 1.8% * | 4.6% * | 5.1% * | 0.4% |
| **Family clinical characteristics** |  |  |  |  |
| General psychopathology (ASR), mean (sd) T-scores | 46.1 (10.6)* | 49.2 (10.4)* | 49.9 (10.4)* | 42.5 (10.1) |
| Maternal mental health service use (%) | 40.9% * | 53.0% * | 58.7% * | 30.1% |
| Paternal mental health service use (%) | 24.6% * | 30.7% * | 33.2% * | 20.2% |
| Maternal mental health hospitalization (%) | 9.0% * | 12.8% * | 15.9% * | 4.5% |
| Paternal mental health hospitalization (%) | 6.8% * | 8.4% * | 10.4% * | 4.0% |
| Maternal history of depression (%) | 32.4% * | 43.2% * | 49.7% * | 22.6% |
| Paternal history of depression (%) | 20.8% * | 25.5% | 30.3% * | 13.2% |
| Maternal history of suicide attempt/death (%) | 6.8% * | 10.0% * | 11.5% * | 2.9% |
| Paternal history of suicide attempt/death (%) | 4.8% * | 5.8% * | 8.7% * | 2.1% |
| Maternal alcohol/substance use during pregnancy (%) | 13.0% * | 13.5% * | 14.1% * | 8.2% |
| * Denotes group differences at $p_{FDR} < 0.05$ between the never-suicidal group and each of the groups endorsing suicidality.<br>ADHD, Attention-deficit/hyperactivity disorder; ASR, Adult Self-Report; CBCL, Child Behavior Checklist; CD, Conduct disorder; ODD, Oppositional defiant disorder; PTSD, Posttraumatic stress disorder; sd, standard deviation.<br>Definitions of each variable can be found in Supplementary Table S1. | | | | |

### Differences and equivalence of neuroimaging data

**Supplementary eFigure 1** shows the sample size of the groups for each imaging modality analyzed. For each modality, we provide the combined results of applying traditional null-hypothesis Welch’s t-tests and Equivalence test, after applying FDR-correction for multiple comparisons. The distribution of results is depicted in **Figure 1**.

Results at corrected and uncorrected level for all modalities are summarized in **Supplementary eTables 3 and 4**, along with brain measures, if any, that showed to be statistically different *and* not statistically equivalent across two or more group comparisons.

### Brain structural imaging

Among the 86 regions examined, only the left bank of the superior temporal sulcus was found to be significantly thinner in the caregiver-reported suicidality group than in the never-suicidal group after applying FDR-correction (ES=-0.17, 95%CI -0.26, -0.08, *p*_FDR_=0.019) (**Figures 1-2, Supplementary eTables 5-7**). In addition, based on our prespecified bounds of ±0.15, this effect was large enough to be considered meaningful.

**Figure 2.**
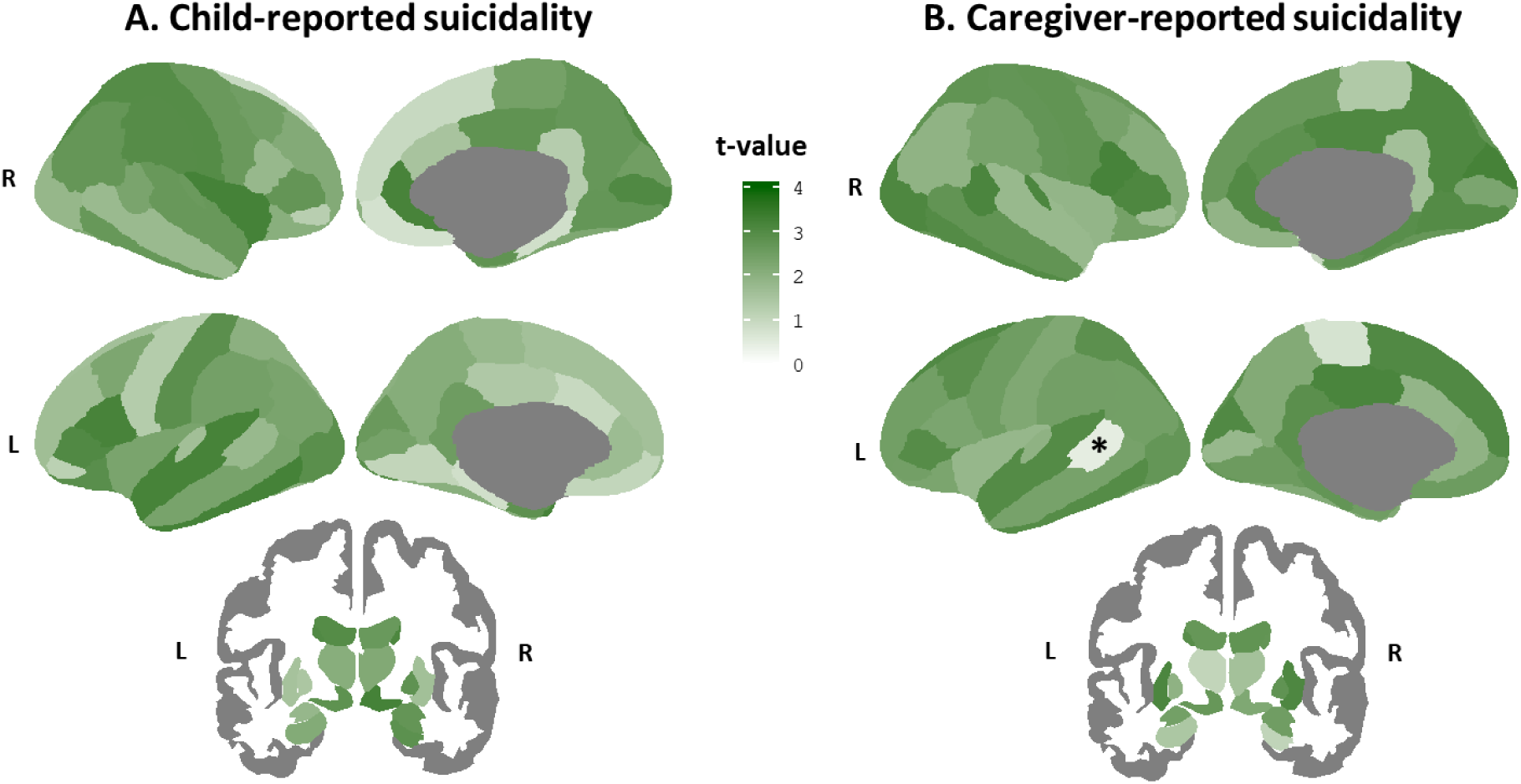
Equivalence testing for mean differences in brain cortical thickness and subcortical volumes relating to suicidality. Distribution of t-values from Equivalence tests comparing the regional means between the never-suicidal group (n=5,381) and the child-reported suicidality group (n=525) (Panel A) and the caregiver-reported suicidality group (n=482) (Panel B); Higher t-values (i.e., darker green) suggest equivalence between groups. *Only the left bank of the temporal sulcus in the caregiver-reported suicidality analysis showed to be statistically different and not statistically equivalent after FDR-correction.

All the remaining regions showed to be not statistically different (all *p*_FDR_>0.05). Of these, most regions were statistically equivalent (i.e., ES were practically zero) for the child-reported suicidality comparison (62 regions [72.1%], ES range=-0.07, 0.07) and for the caregiver-reported suicidality comparison (76 regions [88.4%], ES range=-0.06, 0.07). In contrast, for the concordantly-reported suicidality comparisons, all regions were found to be not statistically equivalent (i.e., ES 90%CI included zero and overlapped with at least one of the |0.15| bounds) with ES ranging -0.23 to 0.23 (**Figure 1, Supplementary eFigures 2-7**).

### Resting-state functional imaging

Among the 306 functional connectivity measures, none showed to be statistically different after applying FDR-correction (all *p*_FDR_>0.05) (**Figure 1, Supplementary eTables 8-10**). In addition, most functional connectivity measures were statistically equivalent (i.e., ES were practically zero) for the child-reported suicidality comparison (170 [55.6%], ES range=-0.04, 0.04). In contrast, for the caregiver- and concordantly-reported suicidality comparisons, all functional connectivity measures were found to be not statistically equivalent (i.e., ES 90%CI included zero and overlapped with at least one of the |0.15| bounds) with ES ranging -0.18 to 0.20, and -0.34 to 0.28, respectively (**Supplementary eFigures 8-10**).

### Task-based functional imaging

Results of Welch’s t-test and Equivalence tests for each of the tasks and contrasts examined are shown in **Figure 1, Figure 3, Supplementary eTables 11-34**, and **Supplementary eFigures 11-60**. Briefly, among the 167 ROI mean activations examined for each of the 3 tasks and 8 contrasts, none showed to be statistically different after applying FDR-correction (all *p*_FDR_>0.05).

**Figure 3.**
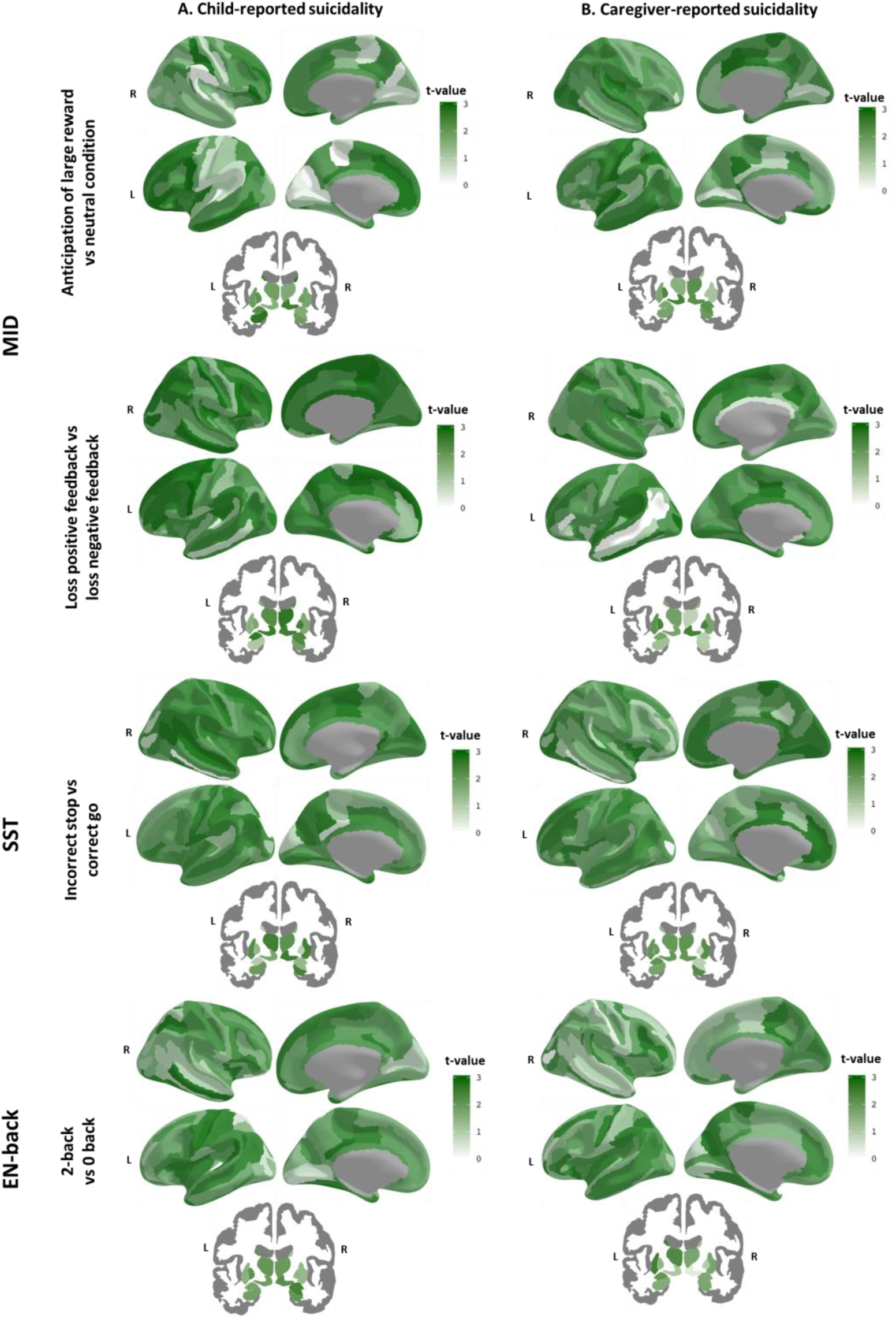
Equivalence testing for mean differences in functional activation during the MID task, the SST and EN-back task relating to suicidality. Distribution of absolute t-values from Equivalence tests comparing the regional means between the never-suicidal group and the child-reported suicidality group (Panel A) and the caregiver-reported suicidality group (Panel B); Higher t-values (i.e., darker green) suggest equivalence between groups. No statistical differences were found after applying FDR-correction for multiple comparisons. The remaining contrasts are depicted in Supplementary eFigure 60.

Of the ROI mean activations, the number of statistically equivalent measures ranged between 0-126 (0%-75.4%, ES range=-0.05, 0.05) for the child-reported suicidality comparison, 0-116 (0%-69.5%, ES range=-0.04, 0.04) for the caregiver-reported suicidality comparison and were none for the concordantly-reported suicidality comparison (**Figure 1, Figure 3, Supplementary eFigure 60**). No evidence of equivalence was found for 41-167 (24.6%-100%, ES range=-0.17, 0.16), 51-167 (30.5%-100%, ES range=-0.17, 0.20), and 167 (100%, ES range=-0.34, 0.25) of ROI mean activations for child-, caregiver-, and concordantly-reported suicidality comparisons, respectively. The MID task showed the higher rates of ROI activations that were statistically equivalent, followed by the SST, and EN-back (**Figure 1, Figure 3, Supplementary eFigure 60**).

### Predictive value of neuroimaging data

Overall, observed ES were small, especially for child- and caregiver-reported suicidality analyses. Maximum ES for child-, caregiver- and concordant-analyses were |0.17|, |0.20|, and |0.34|, respectively (**Supplementary eTable 35)**. Based on lowest and highest 90%CI bounds, all results would have been statistically equivalent if thresholds were |0.29|, |0.33|, and |0.56| for child-, caregiver, and concordantly-reported suicidality comparisons, respectively.

For child- and caregiver-suicidality comparisons, only 23 tests (0.67%) resulted in an ES equal or over our smallest ES of interest (*d*≥|0.15|) (**Figure 4, Supplementary eTable 36)**. These included lower thickness of the left bank superior temporal sulcus, aberrant connectivity of the default and cingulo-parietal network with hippocampus and other subcortical areas, and aberrant task-elicited activation of frontal, temporal, and parieto-occipital areas, and insula. The AUPRC of these observed ES ranged 0.07 to 0.10. Based on the prevalence of suicidality on child- and caregiver-reports in our sample (∼8.5%), these can be considered random classifiers. The AUPRC of the largest ES, found in the sensorimotor mouth-visual area connectivity in the concordant group analysis (*d*=0.34, 95%CI -0.55, -0.12) was 0.02.

**Figure 4.**
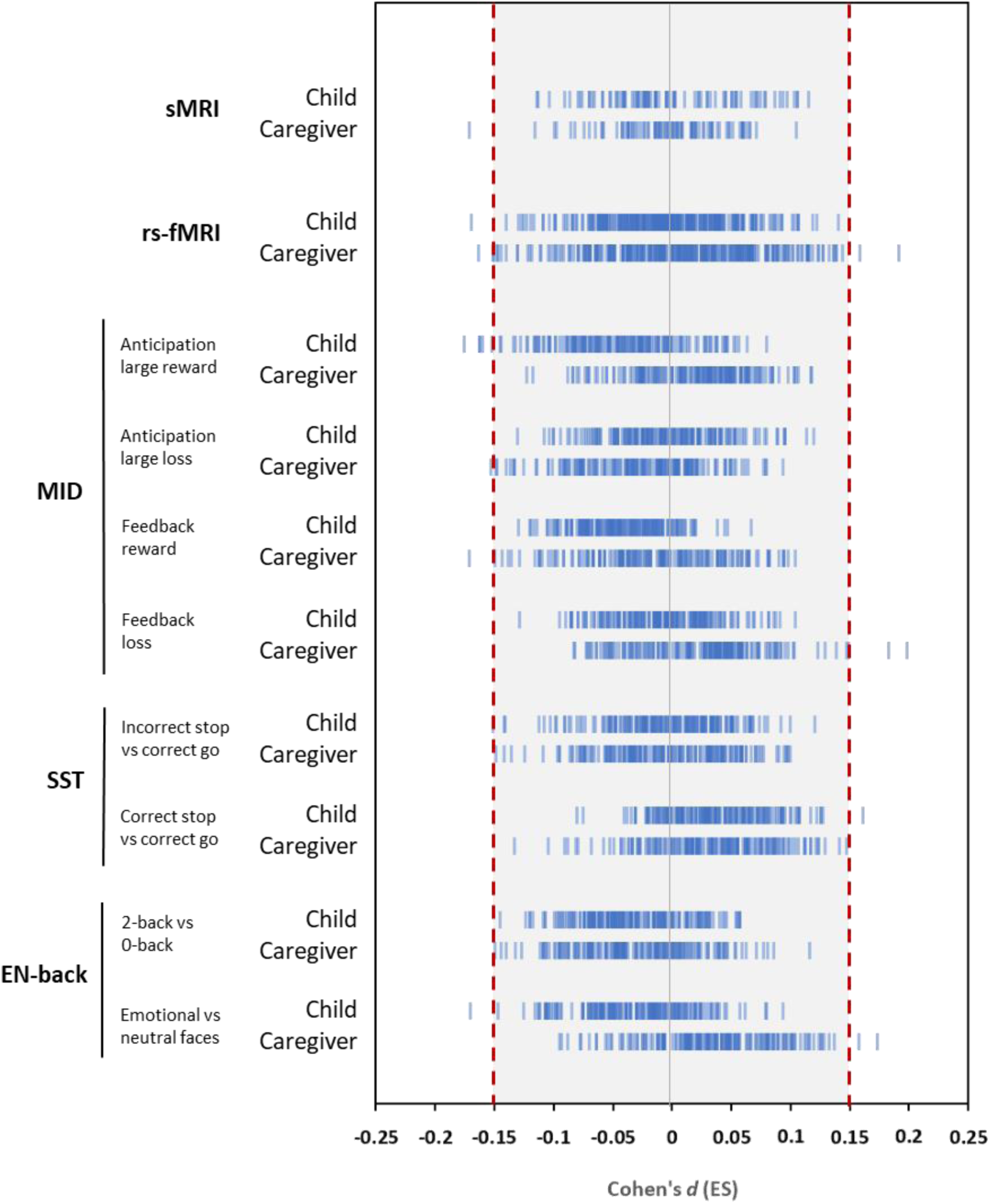
Observed effect sizes (ES) of mean differences for each imaging modality by suicidality group comparison. For each informant, structural MRI examined 86 regions, resting-state fMRI examined 306 connectivity indices, and task-based fMRI examined activations in 167 regions. Blue individual lines represent ES of group mean differences for a region or connectivity index. Shaded area represents ES lower than the prespecified smallest effect size of interest (SESOI) of *d*=|0.15|. sMRI, structural MRI. rs-fMRI, resting-state fMRI. MID, Monetary incentive delay task. SST, Stop signal task. EN-back, Emotional n-back task.

## Discussion

In a large US population-based sample of school-aged children we found that suicidality endorsement was associated with higher levels of psychopathology and social adversity. However, over the 5,000 tests performed to examine differences in structural MRI and resting-state and task-based fMRI, only one survived correction, in which suicidality was associated with thinner left bank of the superior temporal sulcus. Nevertheless, effect sizes were very small, and their ability to predict cases with suicidality was not better than random selection.

The rate of reported suicidality in our sample was in line with rates found in pre-pubertal and school-aged children ^50,51^, which is lower than in community samples of adolescents and young adults ^5-8,52^. Child and caregiver reports of suicidality were not consistent, which is a common observation in adolescents and young adults in whom non-disclosure might involve concerns about stigmatization, difficulties in communication and unavailability of social and family support ^53^. Regardless of informant, though, suicidality was associated with higher psychosocial adversity and clinical correlates thus replicating a number of studies ^5-11,13,14,30,52^.

In terms of neuroimaging correlates, at uncorrected level, we found several regions associated with suicidality not consistently reported in the literature ^18^; and those regions that we found that have been reported (e.g. aberrant thickness in medial orbitofrontal gyrus, aberrant connectivity in the default mode and salience networks, or aberrant task-elicited activations in temporal lobe and insula) differed in directionality or specific regions involved ^18,38,39,54-57^. Moreover, after FDR-correction, we only found a thinner left bank of the superior temporal sulcus in the caregiver-reported suicidality analysis. Similar findings have been found in adults with schizophrenia ^58^. The superior temporal region is part of a neural network involved in inhibitory control and emotion processing in social contexts and has been associated with lethality of attempts and impulsivity ^59^.

Regardless of differences, and based on our prespecified conservative bound of |0.15|, we showed that around half of the group means for child-reported suicidality comparisons (∼48%), and a fifth for parent-reported suicidality comparisons (∼22%) were equivalent (i.e., not a meaningful effect); these would have been nearly 100% equivalent with a prespecified bound of |0.30|, which is still small. In the case of the concordant group, all observed ES of mean differences were not statistically equivalent (i.e., meaningful effects). In these cases, where there is no difference, but effects are not statistically equivalent, there is insufficient data to draw conclusions. With our conservative SESOI of |0.15| the equivalence bounds became narrower and the concordant group should have had a larger sample size in order to obtain a sufficiently narrow confidence interval to conclude that the observed ES were statistically equivalent (i.e., not a meaningful effect).

Of note, observed ES were relatively small for all regions and connectivity indices tested (*d*<|0.30|) in line with studies conducted in large samples ^23^, even within the concordant group. Small ES can still be clinically relevant if they can predict clinical outcomes, treatment response, or point to mechanistic pathways of disease ^60^. We therefore examined the predictive value of the largest ES in our sample. We found that these were not better at predicting suicidal cases than what one would get by selecting cases randomly from the population. This is important because, ironically, the shift from studying psychosocial risk factors to neurobiological biomarkers of suicidality was partly motivated by the poor sensitivity of the former in predicting suicide ^15-17^. While the pattern of increasing suicide rates in young people does not give signs of stopping, it is yet not clear whether this change in focus of study is providing us with any benefit, especially given the cost of neuroimaging studies. The aim was to improve identification and prevention of suicidality; however, to date, the evidence is still weak for this purpose due to small sample sizes, heterogeneity and inconsistency across studies, and, as further shown in this study, small effects sizes with limited predictive value. There is therefore an urgent need to improve the study of neurobiological biomarkers, possibly in conjunction with psychosocial risk factors, using other methodologies such as machine learning ^61,62^. That said, what our results show is that vulnerability to suicidality does not appear to have a “brain signature” with a strong enough effect in school-age children. However, this does not imply that suicidality does not have brain correlates but indicates that such associations, if any, might not discernible using common neuroimaging measures at this age. It is plausible that, as brain organization evolves during the adolescent years, these suicidality correlates become more evident as the brain matures. In that sense, investigation of the longitudinal data from the ABCD cohort when they become available will likely shed some light to these incongruent findings across samples of different ages.

Furthermore, we could combine distinct types of risk, including psychosocial, clinical, and neuroimaging measures, and examine interrelated trajectories across factors that might help us to identifying the shift to more active suicidal behaviors at peak ages such as late adolescence and early adulthood.

### Limitations

Our study has some limitations. Since participants were drawn from the community very few had active suicidal thoughts or behaviors at the time of scanning, and therefore were not necessarily representative of, and comparable to, clinical cases. However, passive ideation has been shown to be associated with significant psychiatric comorbidity and be similar to active ideation in terms of risk factors ^63^, as also shown in this study. In addition, this approach avoids referral biases and might aid the identification of suicidality in community samples. Future waves of ABCD should capture the age-related increase in prevalence of more active suicidal behaviors.

## Conclusions

Vulnerability to suicidality in young children does not appear to have a discrete brain signature when considering commonly-used neuroimaging measures. Moreover, observed effect sizes of imaging correlates of suicidality are small with limited predictive value. There is a great need for improved approaches to the neurobiology of suicide.

## Data Availability

N/A

## Acknowledgement

We thank Dr. Anthony Steven Dick, Associate Professor, Florida International University, for his advice in implementing and interpreting the equivalence tests. We thank Dr. Stephen E. Gilman, Chief of Social and Behavioral Science, National Institute of Child Health and Human Development, for his valuable insight and commentaries on this manuscript.

Data used in the preparation of this article were obtained from the Adolescent Brain Cognitive Development (ABCD) Study (https://abcdstudy.org), held in the NIMH Data Archive (NDA). This is a multisite, longitudinal study designed to recruit more than 10,000 children age 9-10 and follow them over 10 years into early adulthood. The ABCD Study is supported by the National Institutes of Health and additional federal partners under award numbers U01DA041022, U01DA041028, U01DA041048, U01DA041089, U01DA041106, U01DA041117, U01DA041120, U01DA041134, U01DA041148, U01DA041156, U01DA041174, U24DA041123, U24DA041147, U01DA041093, and U01DA041025. A full list of supporters is available at https://abcdstudy.org/federal-partners.html. A listing of participating sites and a complete listing of the study investigators can be found at https://abcdstudy.org/scientists/workgroups/. ABCD consortium investigators designed and implemented the study and/or provided data but did not necessarily participate in analysis or writing of this report. This manuscript reflects the views of the authors and may not reflect the opinions or views of the NIH or ABCD consortium investigators.

